# Neurobehavioral Changes in Fear Generalization in Clinical High Risk for Psychosis

**DOI:** 10.64898/2026.01.14.26344102

**Authors:** Jacqueline A. Clauss, Baktash Babadi, Lauri Tuominen, Wisteria Deng, Logan Leathem, Daphne J. Holt

## Abstract

Changes in basic associative learning processes have been identified in several psychiatric conditions. However, it remains unknown whether these findings result from ongoing psychopathology or represent an underlying transdiagnostic risk factor for multiple clinical states. In this study, 72 participants with subclinical psychopathology, 23 of whom met criteria for the clinical high-risk for psychosis transdiagnostic risk syndrome (CHR+), completed a fear conditioning and generalization paradigm while functional magnetic resonance imaging (fMRI) data were collected. During this procedure, participants were presented with face stimuli that were either paired (CS+) or not paired (CS-) with a mild electrical shock, as well as eight face “morphs” (between the CS+ and CS-) tailored to each participant’s perceptual discrimination ability. Following the scan session, participants rated the likelihood that each stimulus had been previously followed by a shock. Fear generalization-related neural responses and the memory ratings were then examined using both categorical and individual-level, psychometric modeling approaches. Expected patterns of fear generalization-related responses were observed in the anterior insula and superior frontal gyrus, and in the memory ratings. The psychometric modeling analysis revealed a significantly greater threshold in responses of the left anterior insula, representing a wider fear generalization curve, in the CHR+, compared to the control, group. Moreover, across the whole sample, symptoms of anxiety were associated with a wider fear generalization threshold. Thus, these findings suggest that specific features of one basic associative memory process, fear generalization, may be linked to CHR status and transdiagnostic risk for psychiatric illness.

## Introduction

Adolescents and young adults who meet criteria for the Clinical High-Risk for Psychosis syndrome present with a broad range of psychiatric symptoms (1,2). Although Clinical High-Risk for Psychosis is defined by the presence of attenuated, subclinical positive (psychotic) symptoms, affective symptoms,and the negative symptoms of psychotic disorders, such as anhedonia, diminished affective expression and verbal output, are also often present (1,2).

Given this complex clinical picture, Clinical High-Risk for Psychosis is now typically conceptualized as a transdiagnostic risk syndrome (3,4) which confers a 20-fold increased risk for developing psychotic disorders (5) and a 3-4-fold increased risk for mood and anxiety disorders (6), when compared to general population rates. Understanding the neural mechanisms underlying the different symptom dimensions manifested in Clinical High-Risk for Psychosis may facilitate the development of individualized, mechanism-based interventions for the syndrome.

One basic category of processes that has been implicated in a range of psychiatric disorders is associative learning, which involves the pairing and subsequent association of two unrelated stimuli. For example, in studies using Pavlovian fear or aversive conditioning paradigms, individuals with anxiety are more likely than healthy controls to exhibit a fear response to a stimulus that is perceptually similar to a stimulus that was previously associated with an aversive experience such as a shock, i.e., showing greater *generalization* of conditioned fear responses (7,8). In addition, prior studies have also linked psychotic symptoms to abnormalities in associative memory processes, including prediction error signaling (9), reward and aversive conditioning (10–13), and extinction memory (14,15). Also, the negative symptoms of psychotic disorders have been linked to impairments in the acquisition and retrieval of associative memory traces (16) and in fear generalization (17), suggesting that both the positive and negative symptoms of psychotic disorders may be linked to changes in associative learning.

One possible explanation for this wide range of prior findings is that a specific, common component or stage of associative learning is disrupted across samples and tasks. Stimulus generalization is one such simple, fundamental process, defined as the ability to rapidly retrieve information about a learned stimulus and use this information to respond to similar, but novel stimuli (18). Generalization is critical for rapidly determining whether to approach or avoid a novel stimulus, and the generalization of previously learned responses to aversive stimuli (“fear generalization”) is an evolutionarily adaptive, automatic process for prompt detection of potential environmental threats (18).

Generalization relies on both memory retrieval and perceptual discrimination processes (19,20); thus experimental paradigms measuring generalization must account for these component mechanisms. A prior study which controlled for the perceptual discrimination ability of the participants found evidence for diminished generalization of a conditioned association in individuals with psychotic disorders (17). Moreover, decreased generalization in the individuals with psychotic disorders was correlated with negative symptom severity. This finding suggests that some of the impairments in motivation and emotional expression (negative symptoms) observed in psychotic disorders could be due to a reduced ability to quickly assess novel information based on stored associations (21–23). In contrast, there is evidence that anxiety symptoms may involve over-generalization of learned associations to novel stimuli (7,18,24,25). Thus, taken together, prior studies suggest that several common symptoms of psychopathology (the positive and negative symptoms of psychotic disorders and anxiety) may involve disruptions of components of stimulus generalization.

The network of brain regions involved in fear generalization includes the anterior insula and dorsomedial prefrontal cortex (17,21), both of which are components of the salience network. During Pavlovian fear conditioning and generalization paradigms, these regions show activation to stimuli associated with an aversive stimulus (the CS+) and to “generalization” stimuli that are more similar to the CS+ than the control stimulus (CS-). In contrast, regions of the default mode network, such as the posterior cingulate cortex and angular gyrus, show larger responses to the CS-compared to the CS+ and CS+-like generalization stimuli (17,21). Compared to healthy control subjects, individuals with psychotic disorders show diminished responses of the salience network and greater responses of the default mode network to CS+-like stimuli during fear generalization (17).

One remaining unanswered question is whether impairments in fear generalization are present at earlier or less differentiated stages of psychopathology, such as in individuals meeting Clinical High-Risk for Psychosis criteria. To address this question, in this study we measured fear generalization in non-help seeking young adults who met criteria for Clinical High-Risk for Psychosis, comparing them to demographically matched control subjects. We used a previously validated fear generalization paradigm that accounts for individual differences in perceptual discrimination (17,20,21) and measured neural responses and shock expectancy ratings (reflecting memory retrieval) during and following the experimental paradigm, respectively.

Previous studies of fear generalization in psychiatric disorders have relied on group-averaged results, which may obscure variation across individuals that is important for understanding clinically heterogeneous syndromes such as Clinical High-Risk for Psychosis. Thus, in the analyses of this study, in addition to conventional between-group comparisons, we also applied a computational modeling approach to the data, to quantify the degree of under-or over-generalization for each individual participant (26). Each participant’s generalization pattern was summarized by variables reflecting the height and width of the generalization curve (27); a lower-than-expected maximum of the curve is thought to reflect impaired retrieval of the learned association, whereas a widened curve may indicate increased fear generalization to novel, benign stimuli.

Therefore, in this study, we tested the prediction that, as observed in psychotic disorders, Clinical High-Risk for Psychosis is associated with diminished fear generalization (poor retrieval of the CS+ memory trace and reduced recruitment of the network of brain regions involved in fear generalization) and this impairment is linked to levels of negative symptoms. In addition, in light of prior evidence for poor pattern separation or over-generalization in association with positive symptoms and anxiety (17,28–30), we predicted that broader generalization gradients would be linked to these symptom clusters.

## Materials and Methods

### Participants

Participants were recruited from three local universities using in-person mental health screenings as part of an ongoing longitudinal study of young adults with transdiagnostic risk factors for developing psychiatric illnesses (31–36). Seventy-two participants (65.6% female, mean age = 19.6) were enrolled in this neuroimaging study (**Table 1**). Participants were eligible for this study if they were deemed to be at transdiagnostic risk for a psychiatric illness based on having an elevated score on a measure of psychotic experiences, primarily delusional beliefs, the Peters et al. Delusions Inventory (PDI; PDI > 7) (37) or a measure of symptoms of depression, the Beck Depression Inventory (BDI; BDI > 5) (38). Some participants with low PDI and BDI scores (PDI < 8 and BDI < 6) were also enrolled in order to increase dimensional variance in the measures across the sample (31–36). All participants were proficient in English and had normal or corrected-to-normal vision based on Snellen acuity, and all participants provided written informed consent in accordance with the guidelines of the Partners Healthcare Institutional Review Board.

**Table 1.** Participant characteristics.

| Measure | CHR+ Group<br>(N = 22) | CHR- Group<br>(N = 59) | t-score | df | p-value |
| --- | --- | --- | --- | --- | --- |
| | Mean ± SD | Mean ± SD | | | |
| Age (years) at baseline | 19.6 ± 0.14 | 19.6 ± 0.18 | 0.29 | 46.8 | 0.77 |
| | N (%) | N (%) | $\chi^2$ | | p-value |
| Gender (% Male) | 11 (50%) | 14 (24%) | 1.5 |  | 0.22 |
| Ethnicity (% Hispanic) | 4 (18%) | 7 (13%) | 0.004 |  | 0.98 |
| Race |  |  | 2.5 |  | 0.65 |
| White | 11 (50%) | 30 (51%) |  |  |  |
| Black | 2 (9%) | 4 (7%) |  |  |  |
| Asian | 8 (36%) | 12 (20%) |  |  |  |
| Mixed Race | 0 (0%) | 2 (3%) |  |  |  |
| Did not respond | 0 (0%) | 1 (2%) |  |  |  |
| | Mean ± SD | Mean ± SD | t-score | df | p-value |
| SIPS Positive Symptoms | 8.5 ± 4.0 | 3.2 ± 2.8 | 5.6 | 30.3 | **< 0.001** |
| SIPS Negative Symptoms | 7.3 ± 5.4 | 2.3 ± 2.9 | 4.0 | 26.6 | **< 0.001** |
| STAI Trait Score | 46.5 ± 9.5 | 37.6 ± 11.0 | 3.4 | 43.5 | **0.001** |

CHR+: clinical high-risk for psychosis group; CHR-: control group, SD: standard deviation; df: degrees of freedom; SIPS: Structured Interview for Psychosis-Risk Syndromes, STAI: State- Trait Anxiety Inventory

Participants also completed a battery of measures of dimensional psychopathology (**Supplemental Methods and Results**).

### Assessments

A trained research coordinator performed the Structured Interview for Psychosis-Risk Syndromes, using the Scale of Psychosis-Risk Symptoms (SIPS/SOPS) (39), to assess whether participants met criteria for the Clinical High-Risk for Psychosis syndrome. Twenty-two participants met criteria for one Clinical High-Risk for Psychosis syndrome, the Attenuated Positive Symptom Syndrome (APSS, “CHR+”) and one participant met the criteria for the Presence of Psychotic Symptoms (POPS) and was thus excluded from analyses. The participants who did not meet Clinical High-Risk for Psychosis syndrome criteria comprised the control group (CHR-, N = 49).

The severity of subthreshold positive symptoms were measured using the SIPS positive symptom scale and subthreshold negative symptoms were measured using the SIPS negative symptom scale (39). In addition, symptoms of anxiety were measured using the State-Trait Anxiety Inventory (STAI), using its “trait” subscale (40).

### Experimental procedures

#### Summary

Following the clinical assessments, participants completed the following procedures: a perceptual discrimination task (see details below), which was followed by a magnetic resonance imaging (MRI) scan session (using a 3T Siemens Prisma MRI scanner with a 64-channel head coil) during which a fear conditioning and generalization paradigm was administered, and skin conductance responses and blood oxygen level dependent (BOLD) data were simultaneously collected. The scan session was then immediately followed by the presentation of the face stimuli viewed during the fear conditioning and generalization paradigm and collection of shock likelihood ratings from participants, in which participants rated how likely each face stimulus viewed during the scanning had been followed by a shock. Lastly, participants completed a second (identical to the first) perceptual discrimination task (which was repeated to ensure stability of the measurements). During the perceptual discrimination task and the fear conditioning and generalization procedures, realistic-appearing, computer-generated face stimuli were presented (17,20,21). The overall design and procedures were similar to those of previous studies of fear generalization (17,20,21,36). Some of the data collected during the fear conditioning phase (testing a different question than the current study) have been previously published (36).

#### Face stimuli of experimental paradigm

Four distinct human faces were created in FaceGen 3.4 (Singular Inversions, Canada) (20) as described previously (21). These faces were assigned to pairs (Pair A and Pair B, see Figure 1). For each participant, one pair was selected and between the two faces in the pair, one was assigned to be the CS+ and the other to be the CS-. The selection of face pairs and assignment of CS+/CS-conditions was counterbalanced and pseudo-randomized across subjects.

**Figure 1.**
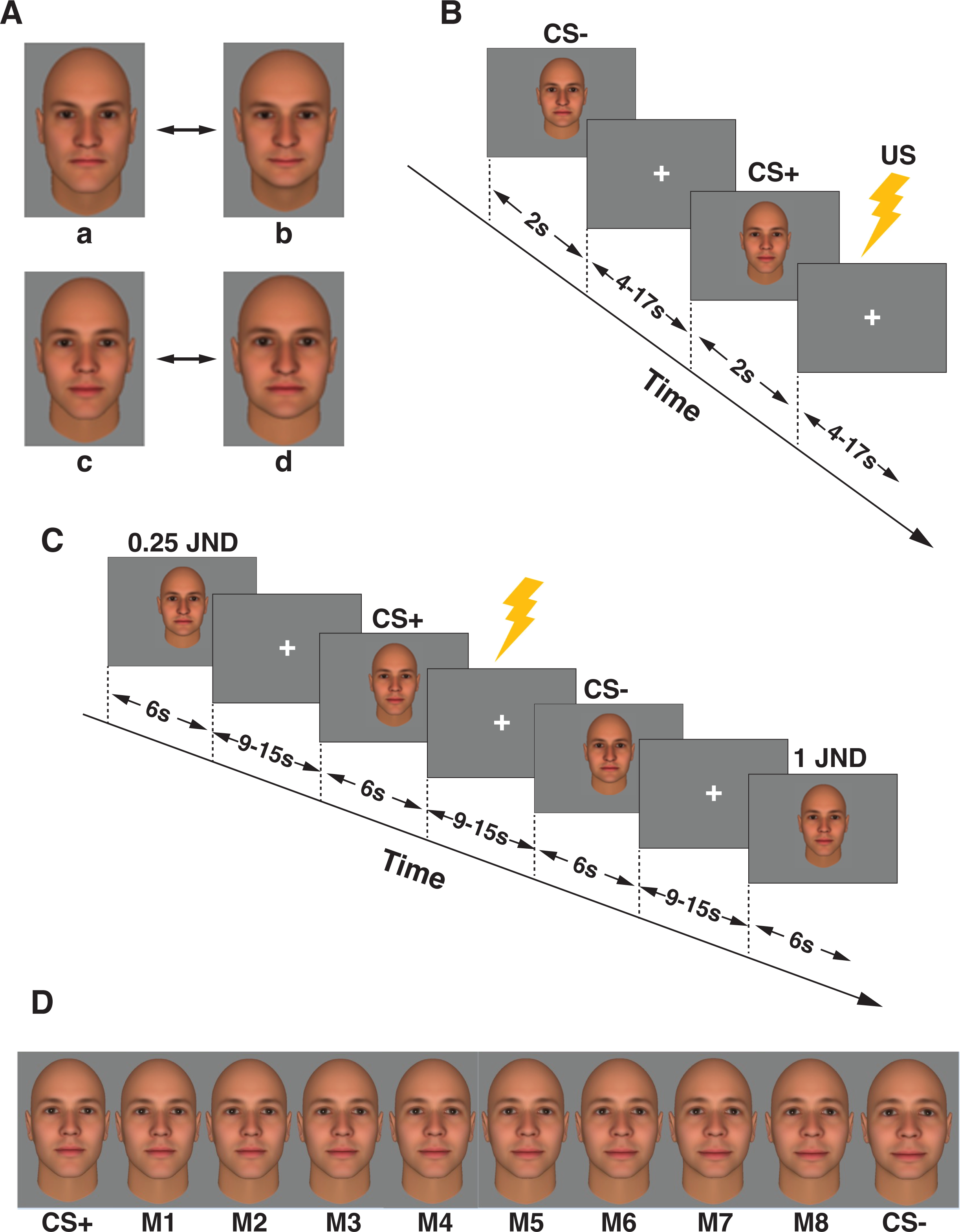
Study design. Participants completed a forced choice perceptual discrimination task during which the Just Noticeable Difference (JND) between A) two face pairs (randomly chosen between A/B and C/D; one face was assigned to be the CS+ and one to be the CS-.) Next, participants underwent B) fear conditioning while fMRI data were collected, during which the CS+ and CS-were presented for 2s each and the CS+ was followed by a shock for 8 of the 13 CS+ trials (61.5%). Time between stimuli ranged from 4-17s jittered. C) Finally, participants completed a fear generalization task while fMRI data were collected, during which the CS+, CS-and morphs ranging from the CS+/CS-to 1 JND, with intermediate morphs between the CS+ and CS-(see Methods), were presented. During the fear generalization phase, the CS+ was always reinforced by a shock, and each stimulus was presented for 6s and the interstimulus interval was 9-15s. D) An example of one individual subject’s fear generalization stimuli is shown, ranging from the CS+ to the CS-with eight morphs in between.

#### Forced-choice perceptual discrimination task

To define each participant’s perceptual threshold, all participants completed a forced-choice perceptual discrimination task (21) (**Supplemental Methods**). The morph that they could differentiate from the CS+ at a 75% accuracy rate was the morph stimulus corresponding to the “just noticeable difference” (JND) threshold for the CS+ (the JND+). The forced-choice task was then repeated for the CS-to define a JND for the CS-(the JND-). This task was administered twice, before and after the MRI scan and shock likelihood ratings.

#### Fear conditioning and generalization paradigm

Participants underwent a Pavlovian fear-conditioning (41) and generalization procedure while fMRI data and skin conductance responses were collected (**Figure 1, Supplemental Methods**). To ensure that participants were attending to the stimuli, participants performed a button-press task during the procedure. Specifically, 30% of the face stimuli appeared to “nod”, with each “nod” lasting 500 ms, and the participants were instructed to press a button on an MR-safe button box whenever they observed a nod.

#### Fear conditioning phase

In each trial of the fear conditioning phase, one face was presented for 2s, followed by a 4-17s inter-trial interval (ITI), and then the other face was presented for 2s. There were 26 trials in total, 13 with the CS+ face presented first and 13 with the CS-face presented first. For 8 of the 13 trials where the CS+ was presented first (61.5% reinforcement), the CS+ was immediately followed by a 500 ms long electrical shock (the unconditioned stimulus; US) administered to the shin of the left leg. The intensity of the US ranged from 1.1 to 4 mA across subjects and was set by each subject to be “highly annoying but not painful” (15,42).

#### Fear generalization phase

After the fear conditioning phase, each participant was presented with the CS+, CS-, and eight CS+/CS-face “morphs” whose difference from the CS+ and CS-was determined by the results of the forced-choice discrimination task performed by the same participant, as described above, to span the entire distance between CS+ and CS - (**Figure 1, Supplemental Methods**). Specifically, the eight morph levels were defined as follows:

M1 = 0.125 JND+

M2 = 0.25 JND+

M3 = 1 JND+

M4 = (1 JND+) + 1/3 (JND+ - JND-)

M5 = (1 JND+) + 2/3 (JND+ - JND-)

M6 = 1 JND-

M7 = 0.25 JND-M8 = 0.125 JND-

The CS+ was always followed by a shock during this phase (100% reinforcement) to minimize extinction. The trials were pseudorandomized such that no more than 2 of the same stimuli were presented consecutively.

#### Shock likelihood ratings

Following the scan session, participants were shown the face images corresponding to the CS+, CS-, and the eight morphs in a counterbalanced, pseudorandom order (**Supplemental Methods**). Participants were asked to rate the percent likelihood that each face stimulus had ever been followed by a shock during the preceding scan session.

#### Magnetic resonance imaging data acquisition

MRI data were collected from 71 participants. Of these datasets, 56 fMRI scans passed quality control procedures (16 CHR+, 40 CHR-, 69.6% female, mean age = 19.8 years; **Supplemental Methods**).

#### Skin conductance data acquisition

Skin conductance responses were measured during the fear conditioning and generalization paradigm; however, initial data quality assessments revealed that the data were too variable and unreliable within and across participants to be suitable for the fear generalization data analyses and thus were not analyzed further.

### Analyses

#### Functional MRI data preprocessing

The fMRI data were preprocessed using the standard FSFAST processing pipeline in FreeSurfer (43) version 6.0 (**Supplemental Methods**).

#### Hypotheses testing

Statistical analyses were completed using RStudio statistical software (Version 2023.09.1+494) RStudio in (Version 2023.09.1+494) using packages: readxl, lme4, dplyr, tidyr, purr, stringr, broom, ggplot2, afex, effsize, emmeans, readr, reshape2, Hmisc, afex, lme4. The fMRI analyses focused on 7 regions of interest (ROIs) defined using fMRI data collected from an independent sample of healthy subjects using a similar experimental paradigm. The regions selected were those which showed significant fear generalization-related BOLD responses in the independent sample (21): the bilateral insula, bilateral superior frontal gyrus, bilateral posterior cingulate cortex, and left middle temporal gyrus (**Figure S1**). Average BOLD responses to each condition (CS+, CS-, morphs) were extracted from each individual for these ROIs.

The effects of group and stimulus on responses (BOLD activity or shock likelihood ratings) related to fear generalization were assessed using two-way repeated measures ANOVAs. Significant group-by-stimulus interactions were followed-up with pairwise t-tests comparing the groups at each stimulus type.

#### Definition of fear generalization

Fear generalization (i.e., generalization of the CS+ association) was reflected by the presence of a significantly greater BOLD response, or shock likelihood rating, to the morphs that were perceptually indistinguishable from the CS+ (morphs 1-3), when compared to the CS-. Generalization of the CS-association was reflected by the presence of a significantly lower BOLD response to the morphs that were indistinguishable from the CS-(morphs 6, 7, and 8) compared to the CS+.

#### Psychometric function fitting

For each subject, the BOLD response and the shock likelihood ratings were used to derive the parameters of a psychometric function, which related the data of the subject (activation of the ROIs or ratings) to the morph level of the face stimuli (see **Supplemental Methods** for additional details). Differences in psychometric curve parameters, including the minimum, maximum, slope, and threshold, were compared between groups.

#### Dimensional analyses of associations between fear generalization parameters and symptoms

Correlations between the parameters of the psychometric functions and symptom measures were calculated using Pearson’s R. In these analyses, the SIPS negative and positive symptom subscales and the STAI-Trait were used as the measures of negative symptoms, positive symptoms and anxiety, respectively. Correlations across both groups and within the CHR+ group were measured. Lastly, relationships between negative symptoms, positive symptoms, and anxiety and the significant findings from group-level analyses of the fMRI data and shock likelihood ratings were evaluated in a multiple regression analysis.

## Results

### Demographic and clinical characteristics of the participants

The CHR+ and CHR-group included 22 and 49 participants, respectively. There were no significant differences in age, sex, ethnicity, or race between the two groups (see Table 1). As expected, the CHR+ group had significantly higher levels of self-reported symptoms of psychopathology than the CHR-group (**Table 1, Table S1**), including: higher levels of positive symptoms, negative symptoms, and trait anxiety (all p ≤ 0.001).

### Behavioral performance

Analyses of the data collected during the *forced-choice perceptual discrimination task* revealed that there were no significant differences between the CHR- and CHR+ groups in the pre-scan or post-scan JND thresholds for the CS+ and the CS-, and the pre-scan and post-scan JNDs did not differ within either group (**Table S1**). In addition, analyses of the performance on the attentional task during the scanning showed that there were no significant differences in detection (i.e., missed button presses) between the two groups during either the fear conditioning or generalization phases (**Table S1**).

### Neural responses during fear conditioning

BOLD responses of the seven ROIs during the conditioning phase were measured to confirm that successful acquisition of conditioned fear-related responses occurred in both groups. Both groups showed differential fear conditioning-related responses (CS+ vs. CS-) in all ROIs (**Supplemental Results**).

### Neural responses during fear generalization

Across the full sample (n= 56), there was a significant effect of stimulus (CS+, CS-, or morph) in activation of the bilateral anterior insula, bilateral SFG, left PCC, and left MTG (all p < 0.005, **Table 2**). The main effects of stimulus in the bilateral anterior insula were due significant fear generalization effects in the full sample, i.e., significantly higher responses to the CS+ and morphs 1-3 compared to the CS-(all p < 0.003). In the bilateral SFG, there was a significantly greater response to the CS+ compared to individual morphs (Figure 2, Table S1). Generalization of the CS-to similar morphs was also observed (**Table S1**), except to morph 8, to which there was an elevated response in all ROIs (which was still significantly lower than the response to the CS+). There were no regions showing a significant main effect of group in activation during fear generalization (all p > 0.09).

**Figure 2.**
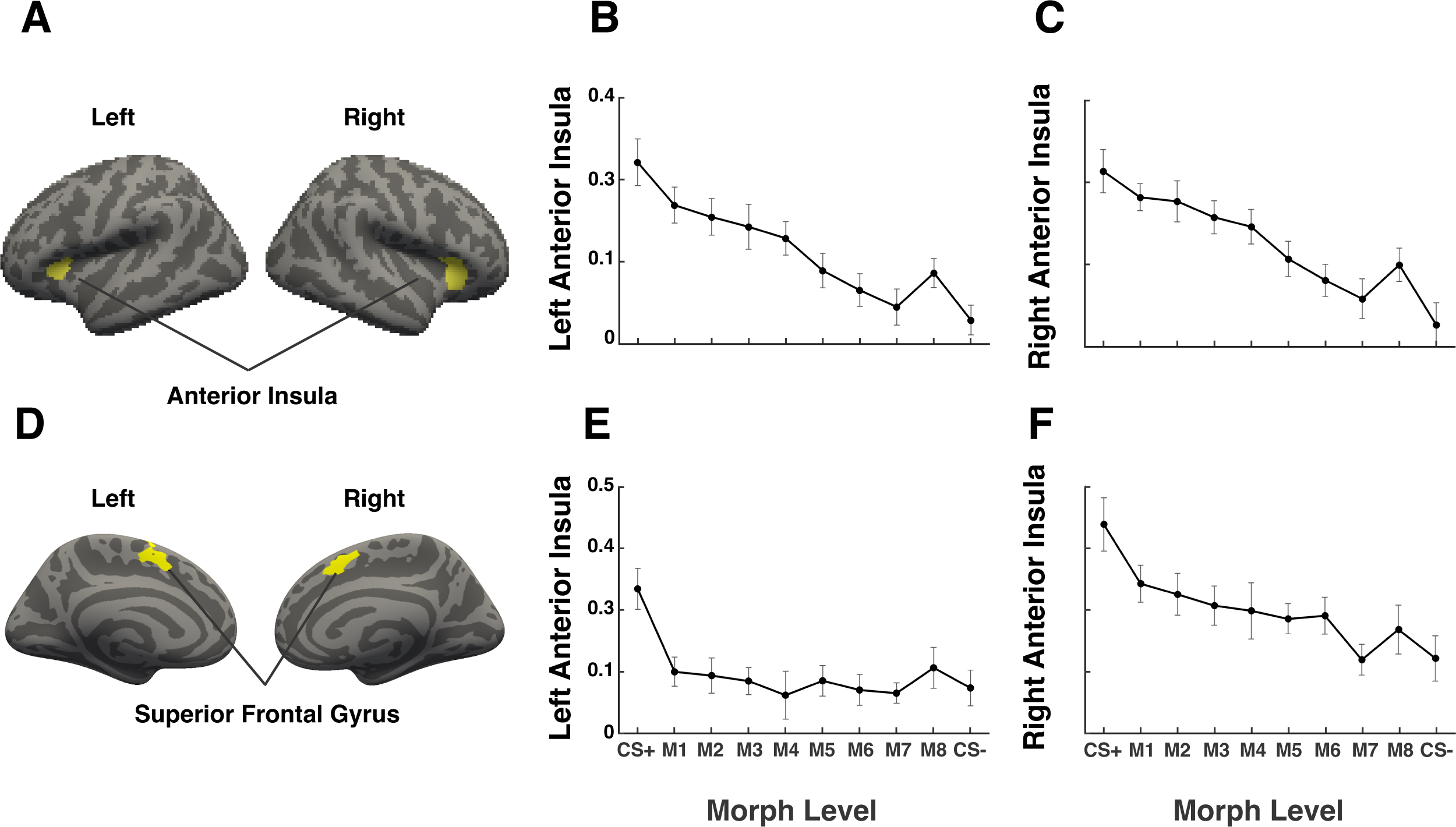
Fear generalization-related BOLD responses in the anterior insula and superior temporal gyrus across the full sample. The anterior insula and superior temporal gyrus regions of interest (ROIs) are shown in panels A and D. There was a significant main effect of stimulus with generalization evident in the responses of the right (B) and left (C) anterior insula ROIs. For the superior frontal gyrus (panels D-F), there was a significant effect of the CS+ compared to individual morphs and an effect of generalization in the CS-to morph 6 and 7, but not to morph 8.

Group x stimulus interactions were observed in the left anterior insula and left SFG responses (**Table 2, Figure S1**), which were due to the significantly lower left anterior insula activation to the CS+ in the CHR+ group, compared to the CHR-group (t(54) = −3.5, p = 0.001) and greater activation in the left SFG to the CS-in the CHR+ group compared to the CHR-group (t(54) = 2.3, p = 0.03).

### Shock likelihood ratings

Following the scan sessions, participants rated the likelihood that each stimulus had been followed by a shock during the fear conditioning and generalization paradigm. There was no significant main effect of group for these ratings (F(69)=0.23, p = 0.64). There was a significant main effect of morph (F(621)=88.8, p < 0.01; Figure 2) and a significant interaction of group x morph (F(621)=2.1, p = 0.025; **Supplemental Results**; **Figure S2**). The main effect of stimulus was due significant generalization effects in the full sample, i.e., higher shock likelihood ratings to the CS+ and morphs 1-3 compared to the CS-(all p < 0.001). There was also evidence for generalization to the CS-, with significantly lower shock likelihood ratings of the morphs that were indistinguishable from the CS-(morphs 6, 7, and 8) compared to the ratings of the CS+.

### Psychometric modeling

We measured the minimum, maximum, slope, and threshold parameters of the fear generalization-related BOLD response functions for the two regions that showed evidence for between-group differences (the anterior insula and SFG) and compared these parameters across the two groups (**Figure 3, Figure S2**). A similar analysis was conducted for the shock likelihood ratings. For the fear generalization responses of the left anterior insula, a significantly higher threshold (p = 0.002) and significantly lower maximum (p < 0.001) was observed in the CHR+ group compared to the CHR-group. A higher generalization threshold suggests that there is less specificity in fear learning (a wider generalization gradient), and the lower maximum indicates diminished recall (i.e., impaired retrieval of the CS+-shock association). There were no significant differences between the two groups in these parameters for the fear generalization responses of the right anterior insula, bilateral SFG, or the shock likelihood ratings.

**Figure 3.**
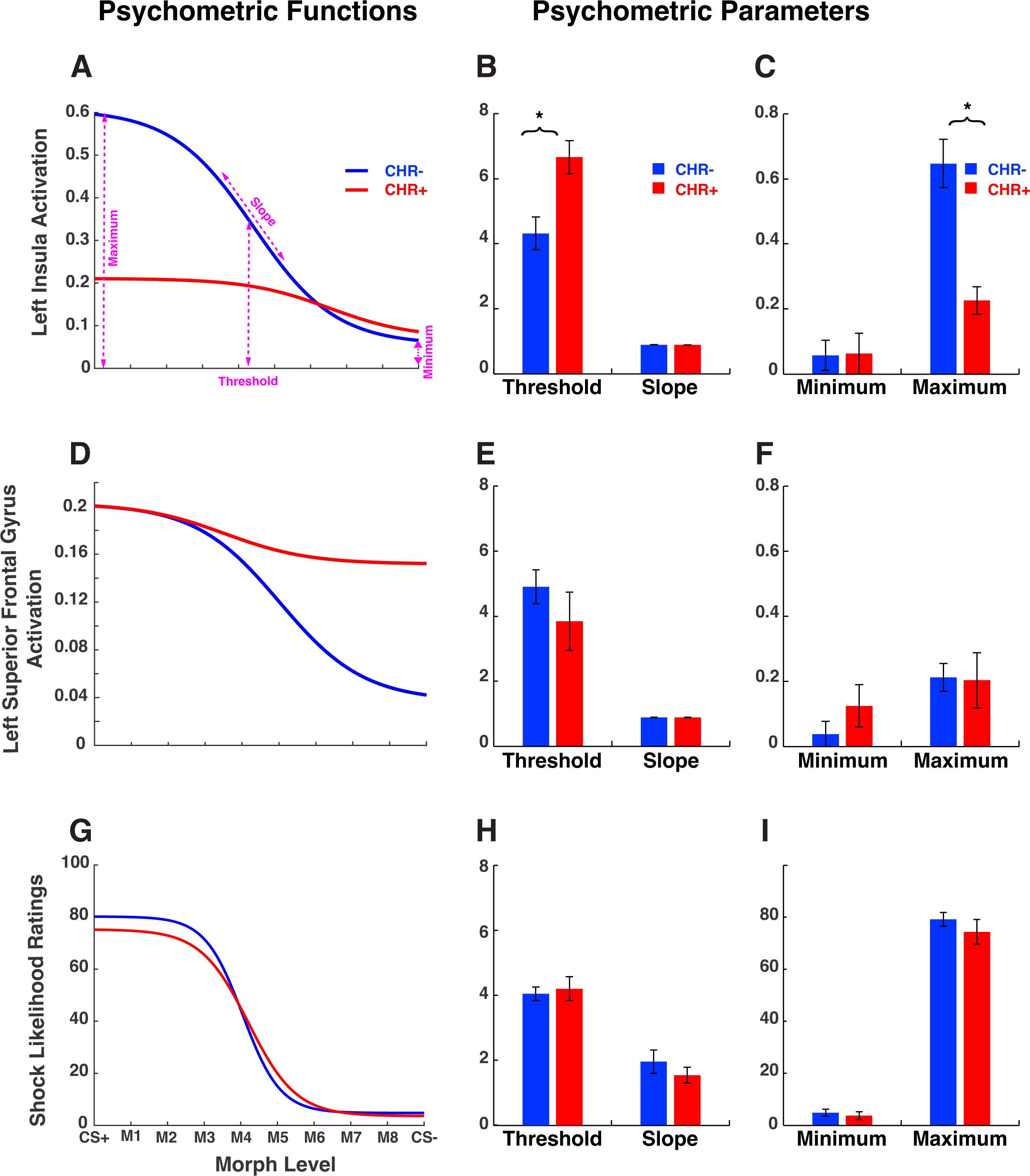
Psychometric modeling results. A) Left anterior insula psychometric function by group. The maximum and threshold of the psychometric function for CHR-group are shown. B) Compared to the CHR-group, the CHR+ group had a significantly larger left anterior insula generalization threshold. C) Compared to the CHR-group, the CHR+ group had a significantly smaller left anterior insula generalization maximum. D) Left superior frontal gyrus psychometric function by group. E) There were no significant differences in left superior frontal gyrus generalization threshold by group or generalization maximum by group. G) Shock likelihood ratings psychometric function by group. There were no significant group differences in H) generalization threshold or I) generalization maximum.

In addition, we tested for relationships between levels of symptoms (positive and negative symptoms and anxiety) and the psychometric function maximum and threshold for both the memory retrieval/shock likelihood ratings and left anterior insula activation within the CHR+ group. Within the CHR+ group, a smaller maximum value of the shock likelihood ratings function in this group was linked to severity of negative symptoms (**Table S4**, r = −0.44, p = 0.04). However, there were no significant correlations between levels of any symptoms and: the generalization threshold for the ratings (all p > 0.10), and the generalization maximum and threshold of the left anterior insula response (all p > 0.2).

### Dimensional analyses (correlations and linear regression) across the full sample

In addition, we measured dimensional relationships across the full sample between levels of the three symptoms of interest (negative symptoms, positive symptoms, and anxiety) and the fear generalization psychometric modeling parameters.

Across the full sample (n= 56), all three symptom types (negative symptoms, positive symptoms, anxiety) were associated with a lower maximum of fear generalization responses across different variables (**Table S5**, *negative symptoms* – left anterior insula response [r = - 0.28, p = 0.04] and shock likelihood ratings [r = −0.31, p = 0.01]; *positive symptoms* – left anterior insula response [r = −0.36, p = 0.01]; *anxiety symptoms* – left anterior insula response [r = −0.35, p = 0.01]). These findings suggest that negative symptoms, positive symptoms, and anxiety are each associated with poor retrieval of associative memory traces during fear generalization, potentially mediated by the left anterior insula and its network.

Additionally, negative symptoms and anxiety were associated with a larger threshold, or a wider fear generalization response (**Table S5**, *negative symptoms*— left anterior insula response [r = 0.32, p = 0.02]; *anxiety symptoms*—left anterior insula response [r = 0.28, p = 0.03] and shock likelihood ratings [r = 0.28, p = 0.02].

As expected, levels of anxiety symptoms, positive symptoms, and negative symptoms were significantly correlated with each other across the entire sample (**Table S5**, anxiety-positive symptoms, r = 0.49, p < 0.01; anxiety-negative symptoms, r = 0.37, p < 0.01; negative symptoms-positive symptoms, r = 0.56, p < 0.01). Therefore, we then conducted a linear regression analysis to assess the relative contributions of each symptom type to the maximum response and threshold of the psychometric functions of the fear generalization-related shock likelihood ratings and the left anterior insula response. There was a significant effect of anxiety (but not the other two symptom types) on the shock likelihood ratings threshold; individuals with more anxiety had a wider fear generalization response (β = 0.04, t = 2.2, p = 0.03, **Figure 4**).

**Figure 4.**
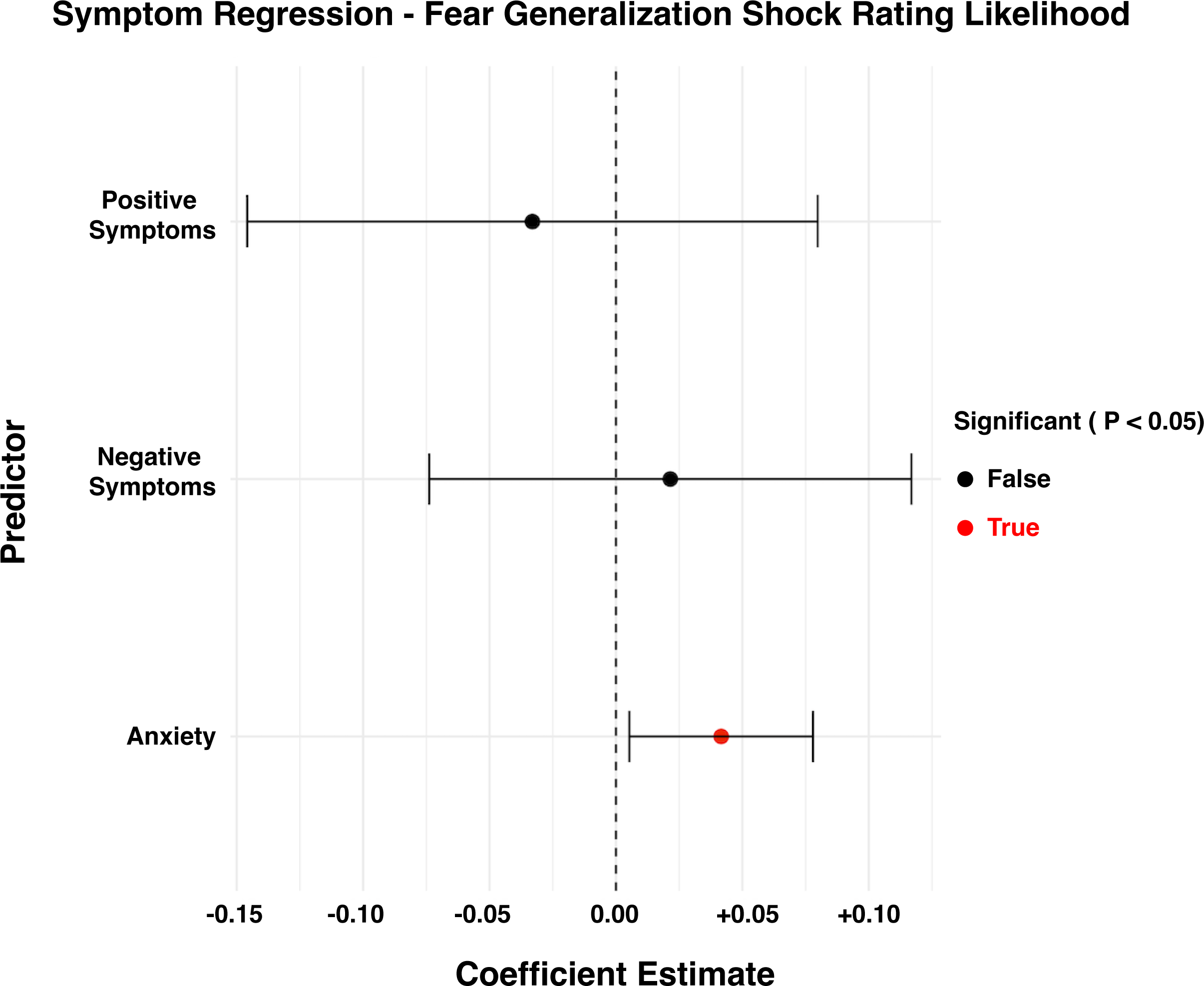
Results of the multiple regression analysis. There was a significant effect of anxiety, but not of the other two symptom types (all p >.10), on the shock likelihood ratings generalization threshold; individuals with higher levels of anxiety had a wider fear generalization response ( = 0.04, t = 2.2, p = 0.03). There were no significant effects by symptom type on the parameters of the left anterior insula generalization response (maximum or threshold).

There were no significant effects of symptom type on the parameters of the left anterior insula response (maximum or threshold).

## Discussion

### Summary of findings

The goal of this study was to determine whether the process of fear generalization is altered in individuals who meet criteria for the Clinical High Risk for Psychosis syndrome. To examine this question, both conventional between-group comparisons and computational modeling of fear generalization-related responses (neural and behavioral) within individual participants were conducted. In this sample, the expected fear generalization-associated responses occurred in the anterior insula and superior frontal gyrus, i.e., the Salience Network (44). In addition, the memory retrieval ratings to the face stimuli viewed during the paradigm revealed the expected pattern of generalization to the morph stimuli that were similar to the CS+ (and to those below the discrimination threshold).

When testing for differences between the CHR+ and control groups, the conventional between-group comparisons revealed little evidence for abnormalities in fear generalization in the CHR+ group. In contrast, the psychometric modeling of the fear generalization functions identified several between-group differences, highlighting the sensitivity of this approach, which quantifies parameters of the fear generalization response on an individual subject basis. In the fear generalization responses of the left anterior insula, a significantly higher threshold (indicating a wider gradient) and a significantly lower maximum was observed in the CHR+, compared to the control group, suggesting that the CHR+ participants showed less specificity in fear memory retrieval (over-generalization) and a lower level of retrieval of the CS+-shock association, respectively. Moreover, levels of negative symptoms were correlated with the maximum values of shock likelihood ratings within the CHR+ group, similar to what was found previously in psychotic disorders (17). Lastly, a linear regression revealed an underlying effect of anxiety on the shock likelihood ratings threshold. Thus, across the full sample, individuals with greater levels of anxiety showed a wider (less specific) fear generalization function, consistent with prior studies of fear generalization in individuals with anxiety disorders (7,8).

Overall, these finding further support prior evidence for abnormalities in associative memory processes, including fear generalization, in psychiatric disorders and provide evidence for this abnormality in a transdiagnostic risk state, Clinical High Risk for Psychosis. An advantage of measuring one specific, low-level component of associative learning, stimulus generalization, in the current study is its fundamental nature (i.e., its relationship to multiple adaptive, more complex functions) and the ability to reliably quantify it within individuals. Future studies could employ this quantitative marker to identify individuals who may be at risk for the development of a range of psychiatric conditions and as a potential objective target for early intervention.

### An individualized approach

A strength of these findings is that they are derived from an experimental paradigm that was tailored to each individual subject, following the identification of the individual’s perceptual threshold (the stimulus morph that they could distinguish from the CS+ stimulus with 75% accuracy, the Just Noticeable Difference). This approach increased the likelihood that each participant would show some fear generalization, potentially increasing the sensitivity of our paradigm to dimensional variation across individuals. Similarly, psychometric curve fitting was conducted initially using each individual participant’s data. Thus, this approach is particularly well-suited to investigating the neurobiological heterogeneity inherent to a clinical category as diverse as Clinical High Risk for Psychosis (2,45) and consistent with other data-driven, clustering approaches that focus less on binary clinically-determined categories and more on the biological and behavioral features linked with psychopathological dimensions (46). These strategies may provide new insights into underlying neurobiological alterations associated with the emergence of psychopathology.

### Comparison to prior studies

The main findings of 1) a significantly higher threshold (indicating a wider gradient) and 2) a significantly lower maximum in the responses of the left anterior insula in the CHR+ group are similar to findings of previous studies of fear generalization conducted in psychiatric disorders. Wider fear generalization curves have been observed in individuals with anxiety disorders and this pattern of findings is consistent with models of anxiety that propose that maladaptive underlying fear generalization processes lead to generalization of fear and avoidance behavioral responses to potential threats in the environment (24,47). Also, one previous study found evidence for a diminished magnitude of fear generalization responses in psychotic disorders (17), similar to our findings of a lower maximum of the fear generalization response function in the CHR+ group. In the prior study of psychotic disorders, the diminished fear generalization responses were correlated with the severity of negative symptoms, similar to the correlation found in the current study between negative symptom levels and the lower maximum of the fear generalization response in the CHR+ participants. Taken together, this pattern of weaker retrieval of the associative memory trace in both populations is consistent with other evidence for abnormalities in various forms of memory retrieval in both psychotic disorders (48) and CHR+ (49). Future studies can also investigate whether a diminished magnitude of fear generalization is linked to specific cognitive impairments in these populations.

Abnormalities in the anterior insula, a component of the Salience Network, have been observed in numerous psychiatric disorders (50–52) and may represent a common, transdiagnostic “hub” showing impaired function across illnesses and early risk states (53–55). Our data further suggest that dimensional variation in anterior insula function, as it affects associative learning and memory, may confer some core vulnerability to psychopathology that is shared across symptom types. This interpretation is consistent with the fundamental role that stimulus generalization plays in cognitive and emotional functions; thus, disruptions of this fundamental process could increase one’s vulnerability to different forms of psychopathology. For example, poor retrieval of associative memory traces when encountering similar stimuli could lead to a failure to protect oneself from potentially harmful experiences, or an inability to consistently approach and engage with potentially beneficial ones. In addition, excessive generalization of a memory trace to stimuli that are distinct from the originally encountered stimulus could lead to maladaptive avoidance of novel experiences. These types of behaviors represent expressions of *transdiagnostic* biases, that when combined with additional deficits or vulnerabilities, could ultimately progress to differentiated, specific conditions (3). However, these hypotheses regarding potential links between transdiagnostic risk for psychiatric illnesses, or later, differentiated psychopathology, and specific features of fear generalization require prospective testing.

In addition, our findings in Clinical High-Risk for Psychosis suggest a testable model for how early symptoms, including negative symptoms and anxiety symptoms specifically, could result in impaired social functioning in this syndrome and associated psychiatric conditions. Individuals with Clinical High-Risk for Psychosis show early deficits in emotion recognition, which have been linked to the later development of psychotic disorders and poorer long-term functioning (56,57). Early impairments in stimulus generalization and functioning of the mediating brain circuitry may contribute to worsening of symptoms (58) and may represent candidate targets for interventions.

### Limitations and future directions

This study has several limitations that should be considered when interpreting its results. The CHR+ participants of this study were not help-seeking and thus may be less likely to develop a psychotic disorder (59) or other psychiatric illnesses. However, it has been well-established that individuals with subthreshold psychotic experiences ranging in severity are at an elevated risk for later functional impairment and developing a broad range of psychiatric illnesses (1). Also, the CHR+ sample of this study was not followed longitudinally, thus the long-term clinical and functional outcomes for these participants remain unknown. Such information, to be collected in future studies, can determine the utility of fear generalization measures for prediction of later onset of psychiatric illnesses, functional impairment and overall prognosis.

Future studies can also determine whether interventions targeting fear generalization impairments could improve longitudinal outcomes for those who meet CHR+ criteria or other transdiagnostic risk states (3,60). For example, emotional resilience-enhancing behavioral interventions for youth may lead to improvements in aspects of emotion regulation (61,62) and emotion recognition (36,63), which have been linked to associative learning functions (64). Also, cognitive remediation may improve memory retrieval and perceptual discrimination (65,66). The use of an quantitative marker of a basic perceptual-memory process such as stimulus generalization, using an individualized approach, could be useful in future trials of preventive interventions, particularly those with a focus on early transdiagnostic stages of risk (60,67). Such studies may lead to the identification of interventions that are specifically beneficial for those with deficits in basic processes involved in the emotional and cognitive functions disrupted in psychiatric illness.

## Data Availability

All data produced in the present study are available upon reasonable request to the authors

## Acknowledgements

The authors are grateful to the participants for their generous time and effort in the study. This work was supported by the Dupont Warren Fellowship of Harvard Psychiatry (JAC), the Louis V. Gerstner Scholar Award (JAC), the Chen Institute Mass General Neuroscience Transformative Scholar Award (JAC), the National Institute of Mental Health (R01MH095904 (DJH), K23MH127508 (BB)).

## Conflict of Interest

The authors declare no competing interests.

## Notes

### Competing Interest Statement

The authors have declared no competing interest.

### Author Declarations

IRB of Partners HEalthcare gave ethical approval for this work.

