## Supplement for "Neurobehavioral Changes in Fear Generalization in Clinical High Risk for Psychosis"

**Supplemental Material**

**Supplemental Methods:**

Dimensional psychopathology: As described in the Methods, the three symptom measures using in the primary analyses were: 1) the SIPS positive symptom subscale, 2) the SIPS negative symptom subscale and 3) the STAI-trait anxiety scale. In addition, participants also completed a number of self-report measures to further capture and explore a range of dimensional psychopathology. Thus the severity of subthreshold positive symptoms was measured using, in addition to the SIPS positive symptom scale (1), the Peters et al Delusion Inventory (PDI) (2), which measures psychotic experiences, primarily delusional ideation, plus the associated distress, preoccupation, and conviction. Negative symptoms were measured using, in addition to the SIPS negative symptom scale (1), the Chapman Social Anhedonia Scale-Revised (SAS-R) and the Chapman Physical Anhedonia Scale (3). The number of social contacts (one indicator of social functioning that may be related to negative symptoms) was measured using the Social Network Index (SNI Contacts) (4). Anxiety symptoms were measured both using the State-Trait Anxiety Inventory-Trait subscale (STAI-T) and the State-Trait Anxiety Inventory-State subscale (STAI-S) (5). Depression was measured using the BDI (6), which assesses symptoms of depression in the past two weeks. The secondary symptom measures were not examined in the primary analyses but merely reported here (see Table S1 below) to corroborate that these symptoms were consistently elevated in the CHR+ (compared to the CHR-) group.

Face stimuli: Four distinct human faces were created in FaceGen 3.4 (Singular Inversions, Canada) (7) as described previously (8). These faces were paired (Pair A and Pair B). For each participant, one pair was selected and between the two faces in the pair, one was selected as the conditioned stimulus (CS+) and one as unconditioned neutral stimulus (CS-). The selection of face pairs and assignment of valence within each pair was counterbalanced and pseudo-randomized across subjects.

Forced-choice perceptual discrimination task: To define a perceptual threshold, all participants participated in a forced-choice perceptual discrimination task as per Tuominen et al. (8). A series of 100 different “morphs” ranging between the CS+ and CS- faces were created. Participants were shown the CS+ face and a face morph and asked to determine if the faces were the same or different. The morph level at which they could differentiate the morph face from the CS+ face with 75% accuracy was the “just noticeable difference” threshold for the CS+ face (JND+). Similarly, the morph level at which they could differentiate the morph face from the CS- face with 75% accuracy was defined as the JND-. The task involved 3 runs of 50 trials each. The CS+ face was presented first for 500ms and then the CS+ face and a morph stimulus were presented side-by-side, and the participant was asked to choose which stimulus they had seen previously. Responses were followed by a 1s inter-trial interval. The position of the CS+ and the morph stimulus were pseudo-randomized. Responses were fit to a Weibull function: y = 1-e(-(x/a)b) in which y was the proportion of correct responses, x was the morph level, and a and b were shape and scale, respectively. The JND value was considered the degree of morph at which the subject achieved 75% accuracy. Eight CS+/CS- hybrids were then chosen based on the JND values for that individual.

Fear conditioning and generalization paradigm: Participants underwent a Pavlovian fear-conditioning and generalization procedure (9) while fMRI data and skin conductance responses (SCRs) were collected. In each trial of the fear conditioning paradigm, one face was presented for 2s, followed by a 4-17s ITI, and then the other face was presented for 2s. There were 26 trials in total, 13 with the CS+ presented first and 13 with the CS- presented first. For 8 of the 13 trials where the CS+ was presented first, the CS+ was immediately followed by a 500 ms-long electrical shock (the unconditioned stimulus; US) applied to the shin of the left leg (61.5% reinforcement). The intensity of the US ranged from 1.1 to 4 mA and was set by each subject to be “highly annoying but not painful” (10,11).

Fear generalization: After the fear conditioning task, participants were presented with the CS+, CS-, and eight hybrid CS+/CS- faces (“morphs”) whose difference from the CS+ was scaled by the results of the forced-choice discrimination task. There were five trials for each stimulus category, totaling 50 trials, and each face was presented for 6 seconds and followed by a 9, 12, or 15-second ITI (see Figure 1). Specifically, the eight morph levels were defined as follows:

M1 = 0.125 JND+

M2 = 0.25 JND+

M3 = 1 JND+

M4 = (1 JND+) + 1/3 (JND+ - JND-)

M5 = (1 JND+) + 2/3 (JND+ - JND-)

M6 = 1 JND-

M7 = 0.25 JND-

M8 = 0.125 JND-

There were five trials for each stimulus category, totaling 50 trials, and each face was presented for 6 seconds and followed by a 9, 12, or 15-second ITI (see Figure 1). The CS+ was always followed by a shock to minimize extinction. The trials were pseudorandomized such that no more than 2 of the same stimuli were presented consecutively.

To promote attention to the stimuli, participants completed a button-press task alongside the trials. Specifically, 30% of the face stimuli appeared to “nod”, with each “nod” lasting 500 ms, and the participants were instructed to press a button on an MR-safe button box whenever they observed a nod.

Explicit ratings: After the scan session (during which participants completed the fear conditioning and generalization task), participants were shown the CS+, CS-, and eight morphs in a counterbalanced, pseudorandom order. The faces were each shown for 6 seconds with a 9 second ITI. Participants were asked to rate the percent likelihood (from 0% to 100%) that the stimulus had ever been followed by a shock during the task.

Magnetic resonance imaging data (MRI) acquisition: MRI data were collected from 71 participants, of which 56 passed quality control procedures (16 CHR+, 40 CHR-, 69.6% female, mean age = 19.8); see below for details.

All MRI data were collected on a 3T Siemens Prisma scanner using a 64-channel head coil (Erlangen, Germany) at the Athinoula A. Martinos Center for Biomedical Imaging. T2*-weighted echo-planar images were collected during the fear conditioning, fear generalization, and resting state (2 mm isotropic, matrix = 64 x 64, 45 slices, TR = 2000 ms, TE = 30 ms, flip angle = 90º). In addition, a T1-weighted 3D scan was collected using an MPRAGE sequence (spatial resolution 1 mm isotropic, matrix = 256 x 256, 176 slices, TR = 2530 ms, TE = 1.64, 3.5, 5.36, and 7.22 ms, flip angle = 7º).

Shock Likelihood Ratings: Following the scan session, participants were shown the face images corresponding to the CS+, CS-, and the eight morphs in a counterbalanced, pseudorandom order. The faces were shown for 6 seconds with a 9 second ITI. Each stimulus was shown twice. Participants were asked to rate the percent likelihood that the stimulus had ever been followed by a shock during the preceding scan session.

Quality control criteria: Volumes with > 1mm of head motion were excluded from analyses. Data of thirteen subjects were excluded due to missing button press data. In addition, two subjects with incomplete imaging data for the fear generalization task were also excluded. Of the 15 participants who were excluded (6 CHR+, 9 CHR-), there were no significant differences in positive symptoms, negative symptoms, or anxiety symptoms between the CHR+ individuals who were excluded versus included and between the CHR- participants who were excluded versus included.

Functional MRI data preprocessing: The fMRI data were preprocessed using the standard FSFAST processing pipeline in FreeSurfer (12) version 6.0. Briefly, functional images were corrected for motion and slice timing and then spatially transformed into a common space (i.e., fsaverage on the cortical surface and MNI152 in the subcortical volume) and then spatially smoothed using a 3D Gaussian kernel (5 mm FWHM). A canonical hemodynamic response function was fitted to the CS+, CS- and morph events in each vertex and voxel. Temporal drift was accounted for by first- and second-degree polynomials in the first-level model. Motion parameters and timepoints with excess motion were used as regressors in the first level model.

Psychometric curve modeling: Using a logistic function, the psychometric function is:

$p\left( x \right)=Min+(Max-Min)/\left( 1+e^{-\left( x-T \right)S} \right)$ ,

where $p(x)$ is the subject’s performance (or BOLD activation), $x$ is the morph level of the face stimuli, $T$ is the threshold, i.e., the morph level yielding half-maximum performance, $S$ governs the slope of the function (larger $S$ leads to a steeper function and a smaller $S$ leads to a flatter function), $Min$is the minimum performance, and $Max$is the minimum performance. A maximum likelihood procedure was used to fit the parameters $T$, $S$, $Min$and $Max$ for each subject in each setting (13).

**Supplemental Results:**

Expected acquisition of fear conditioning-related BOLD responses in the regions-of-interest:

During fear conditioning, there was a significant main effect of stimulus, as defined by a significantly different response to the CS+ versus CS-, for responses of the bilateral angular gyrus, bilateral anterior insula, bilateral posterior cingulate cortex (PCC), bilateral superior frontal gyrus (SFG), and left angular gyrus regions of interest (ROIs; Supplemental Table 2, all p < 0.05). Specifically, as expected, there was significantly greater activation to the CS+ compared to the CS- in the bilateral anterior insula and bilateral SFG (all p < 0.05), whereas in areas of the default mode network, including the left angular gyrus and right PCC, there was significantly greater activation to the CS- compared to the CS+ (all p < 0.05).

There was an overall significant main effect of group for responses of the left anterior insula and bilateral SFG was due to significantly less activation in the CHR+ group compared to the CHR- group (all p < 0.05) in these regions; however, there were no significant group by stimulus interactions. Thus, both groups showed differential fear conditioning-related responses (CS+ vs. CS-) in the ROIs.

Details of fear generalization – related responses observed in the memory (shock likelihood) ratings: The main effect of stimulus was due significant generalization effects in the full sample, i.e., higher shock likelihood ratings to the CS+ and morphs 1-3 compared to the CS- (all p < 0.001). There was also evidence for generalization to the CS- with significantly lower shock likelihood ratings of the morphs that were indistinguishable from the CS- (morphs 6, 7, and 8) compared to the ratings of the CS+. Morph 4 was rated as significantly more likely to be followed by a shock than morphs 5-8 and the CS- (all p < .01). Morph 5 was rated as significantly more likely to be followed by a shock than morph 8 and the CS- (both p < 0.05). Morph 6 was rated as significantly more likely to be followed by a shock than morph 7, morph 8, and the CS- (all p < 0.05). While the group x stimulus interaction was significant, there were no significant between group differences in responses to individual morphs (although there were several trends: morph 1: t(69) = 1.7, p = 0.09, morph 2: t(69) = 1.95, p = 0.06, morph 7: t(69) = -2.0, p = 0.05).

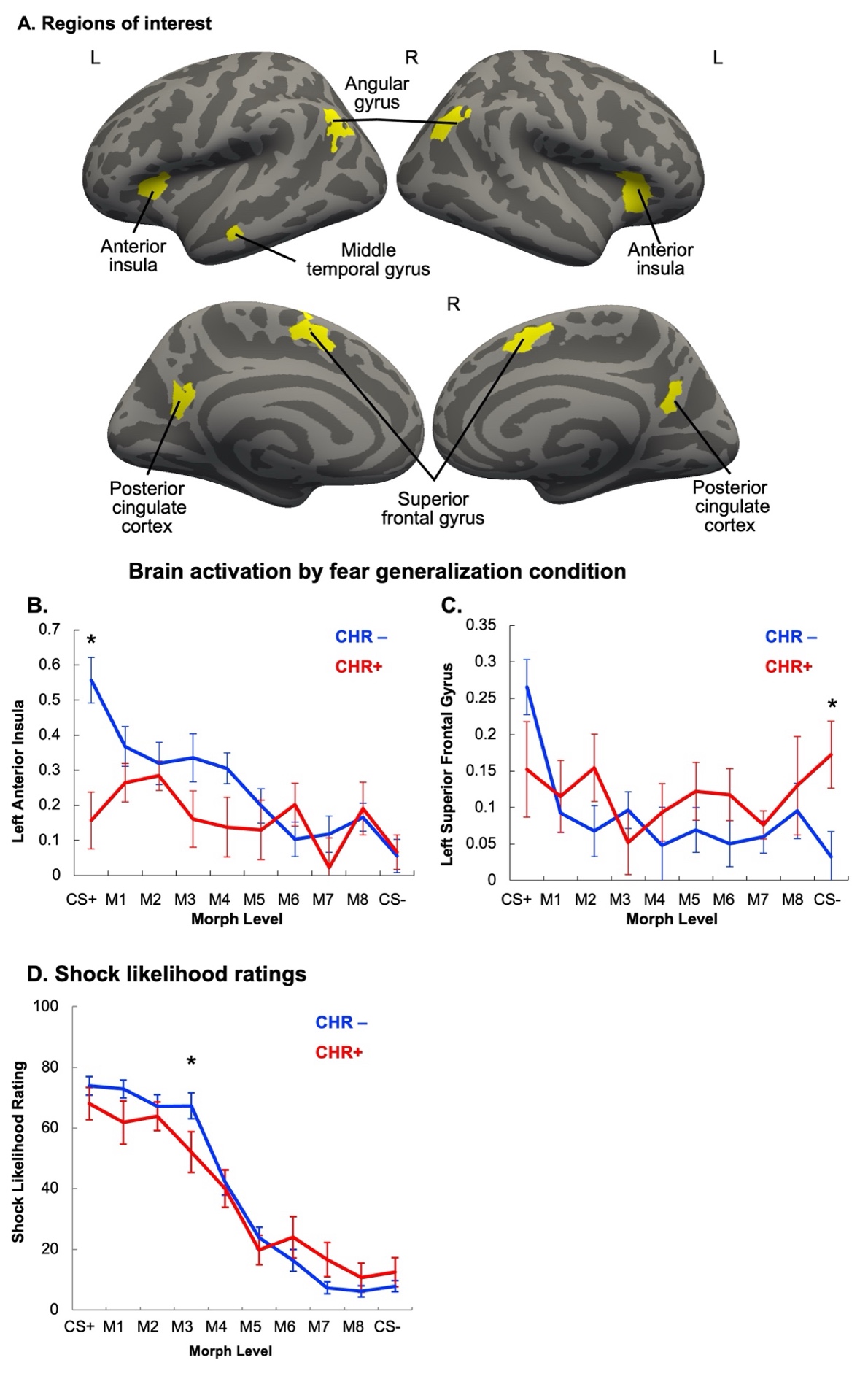
**Figure S1. Regions of Interest and fear generalization-related responses in each group.**

**(A**) The seven regions of interest are displayed on an average brain (“fsaverage”): left and right anterior insula, left and right superior frontal gyrus, left middle temporal gyrus, and left and right posterior cingulate cortex. Group x stimulus interactions were observed in the left anterior insula and left superior frontal gyrus responses, which were due to **(B)** the significantly lower left anterior insula activation to the CS+ in the CHR+ group, compared to the CHR- group (t(54) = -3.5, p = 0.001) and **(C)** the greater activation in the left superior frontal gyrus to the CS- in the CHR+ group compared to the CHR- group (t(54) = 2.3, p = 0.03). **(D)** There was a significant group x stimulus interaction in shock likelihood ratings (F(621)=2.1, p = 0.025). The CHR+ group showed significantly less generalization to morph 3 compared to the CHR- group. * p< .05.

**Figure S2. Psychometric Modeling Results. (A)** Psychometric function of the responses of the left anterior insula by group. **(B)** The CHR+ group had a significantly larger left anterior insula generalization threshold but no difference in slope (p > 0.05). **(C)** There were no significant differences in minima between groups (p > 0.05). The CHR+ group had a significantly smaller maximum of the left anterior insula function. **(D)** Psychometric function of the responses of the left superior frontal gyrus by group. **(E)** There were no significant differences in left superior frontal gyrus threshold or slope by group (all p > 0.05). **(F)** There were no significant differences in left anterior insula minimum or maximum by group (all p > 0.05). **(G)** Psychometric function of the shock likelihood ratings by group. There were no significant group differences in **(H)** slope, threshold **(I)** minimum, or maximum (all p > 0.05).

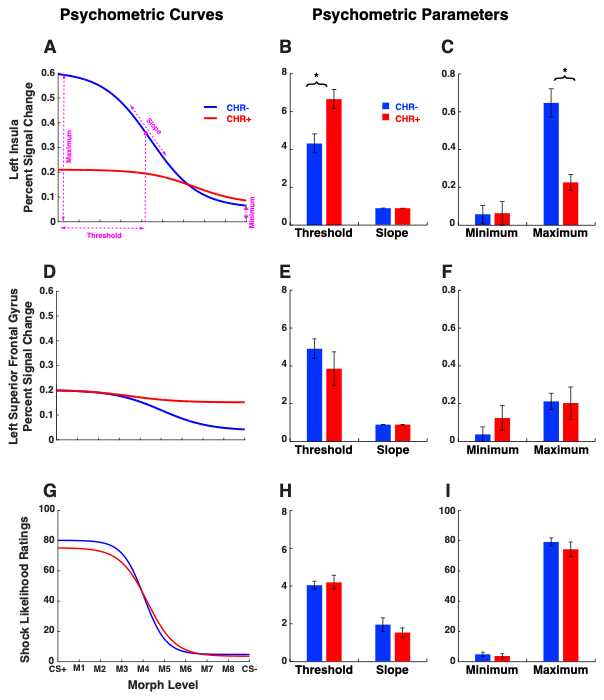

**Table S1. Symptom Measures and Behavioral Data**

|  | **CHR+ Group (N = 22)** | **CHR- Group (N = 59)** |  |  |  |
| --- | --- | --- | --- | --- | --- |
| **Measure** | **Mean ± SD** | **Mean ± SD** | **t-score** | **df** | **p-value** |
| SIPS Positive Symptoms | 8.5 ± 4.0 | 3.2 ± 2.8 | 5.6 | 30.3 | **< 0.001** |
| SIPS Negative Symptoms | 7.3 ± 5.4 | 2.3 ± 2.9 | 4.0 | 26.6 | **< 0.001** |
| BDI Score | 16.2 ± 4.0 | 9.2 ± 3.8 | 4.0 | 36.4 | **0.003** |
| PDI Score | 9.8 ± 4.0 | 5.7 ± 3.8 | 4.0 | 36.4 | **<0.001** |
| STAI Trait Score | 46.5 ± 9.5 | 37.6 ± 11.0 | 3.4 | 43.5 | **0.001** |
| STAI State Score | 38.2 ± 7.6 | 34.2 ± 10.5 | 1.8 | 52.1 | 0.08 |
| SAS | 12.5 ± 7.0 | 6.9 ± 5.5 | 3.2 | 31.3 | **0.003** |
| PAS | 12.3 ± 8.5 | 9.8 ± 9.1 | 1.1 | 40.8 | 0.27 |
| SNI Contacts | 14.5 ± 10.6 | 19.6 ± 9.6 | -1.9 | 35.1 | 0.06 |
| **Behavioral Data** | **Mean ± SD** | **Mean ± SD** | **t-score** | **df** | **p-value** |
| Pre-Task JND CS+ | 27.8 ± 13.0 | 27.3 ± 12.8 | 0.13 | 28.5 | 0.90 |
| Pre-Task JND CS- | 26.4 ± 11.6 | 19.6 ± 10.6 | 0.07 | 26.6 | 0.95 |
| Post-Task JND CS+ | 22.8 ± 11.9 | 30.6 ± 18.1 | -1.95 | 44.2 | 0.06 |
| Post-Task JND CS- | 26.7 ± 13.3 | 29.3 ± 15.9 | -0.64 | 38.5 | 0.52 |
| Missed Button Press – Conditioning | 7.6 ± 6.0 | 9.9 ± 17.8 | -0.64 | 55.6 | 0.47 |
| Missed Button Press – Generalization | 7.5 ± 10.9 | 9.5 ± 11.8 | -0.62 | 29.3 | 0.54 |

CHR+: Clinical High-Risk for psychosis group; CHR-: control group, SD: standard deviation; df: degrees of freedom; SIPS: Structured Interview for Psychosis-Risk Syndromes, BDI: Beck Depression Inventory; PDI: Peters et al., Delusions Inventory; STAI: State-Trait Anxiety Inventory; SAS: Social Anhedonia Scale; PAS: Physical Anhedonia Scale; SNI: Social Network Index; JND: Just Noticeable Difference

**Table S2. Fear conditioning, fear generalization and group results**

| ***Fear conditioning*** | | |  | | | | | |  | | | |  | | |
| --- | --- | --- | --- | --- | --- | --- | --- | --- | --- | --- | --- | --- | --- | --- | --- |
|  | |  | ***Left hemisphere*** | | | | | | | ***Right hemisphere*** | | | | | |
| ***Neural response*** | | | ***F*** | | | ***df*** | | | ***p*** | ***F*** | | | ***df*** | | ***p*** |
| Angular gyrus | | Group | 0.8 | | | 1,54 | | | 0.38 | 0.03 | | | 1,54 | | 0.86 |
|  |  | Condition | 9.5 | | | 1,54 | | | **0.003** | 15.5 | | | 1,54 | | **< 0.001** |
|  |  | Condition x Group | 1.4 | | | 1,54 | | | 0.23 | 0.25 | | | 1,54 | | 0.62 |
| Insula | | Group | 7.4 | | | 1,54 | | | **0.009** | 0.07 | | | 1,54 | | 0.80 |
|  |  | Condition | 31.1 | | | 1,54 | | | **< 0.001** | 38.7 | | | 1,54 | | **< 0.001** |
|  |  | Condition x Group | 2.0 | | | 1,54 | | | 0.16 | 2.1 | | | 1,54 | | 0.15 |
| Posterior cingulate cortex | | Group | 0.004 | | | 1,54 | | | 0.54 | 0.73 | | | 1,54 | | 0.40 |
|  |  | Condition | 4.6 | | | 1,54 | | | **0.04** | 9.6 | | | 1,54 | | **0.003** |
|  |  | Condition x Group | 1.5 | | | 1,54 | | | 0.23 | 1.5 | | | 1,54 | | 0.22 |
| Superior frontal gyrus | | Group | 5.1 | | | 1,54 | | | **0.03** | 4.5 | | | 1,54 | | **0.04** |
|  |  | Condition | 22.8 | | | 1,54 | | | **< 0.001** | 12.8 | | | 1,54 | | **< 0.001** |
|  |  | Condition x Group | 2.3 | | | 1,54 | | | 0.14 | 1.7 | | | 1,54 | | 0.19 |
| Middle temporal gyrus | | Group | 0.38 | | | 1,54 | | | 0.54 |  | | |  | |  |
|  |  | Condition | 7.9 | | | 1,54 | | | **0.007** |  | | |  | |  |
|  |  | Condition x Group | 0.01 | | | 1,54 | | | 0.91 |  | | |  | |  |
| ***Fear generalization*** | | | | | | | | | | | | | | | |
| ***Behavioral response*** | | | ***F*** | | | | | | ***df*** | | | | ***p*** | | |
| Explicit ratings | Group | | .23 | | | | | | 1, 69 | | | | 0.63 | | |
|  | Condition | | 88.8 | | | | | | 9, 621 | | | | **<.001*** | | |
|  | Condition x Group | | 2.13 | | | | | | 9, 621 | | | | **0.026*** | | |
|  | | | *Left hemisphere* | | | | | | | | *Right hemisphere* | | | | |
| ***Neural response*** | | | ***F*** | ***df*** | | | | ***p*** | | | ***F*** | ***df*** | | ***p*** | |
| Angular gyrus | Group | | 0.2 | 1, 54 | | | | 0.63 | | | 0.5 | 1, 54 | | 0.47 | |
|  | Condition | | 1.8 | 9. 486 | | | | 0.06 | | | 1.1 | 9, 486 | | 0.33 | |
|  | Condition x Group | | 0.5 | 9. 486 | | | | 0.84 | | | 0.6 | 9, 486 | | 0.83 | |
| Insula | Group | | 3.0 | 1, 54 | | | | 0.09 | | | 2.4 | 1, 54 | | 0.13 | |
|  | Condition | | 5.6 | 9, 486 | | | | **0.001*** | | | 7.9 | 9, 486 | | **< 0.001*** | |
|  | Condition x Group | | 2.6 | 9, 486 | | | | **0.005*** | | | 1.1 | 9, 486 | | 0.33 | |
| Posterior cingulate cortex | Group | | 0.2 | 1, 54 | | | | 0.67 | | | 0.4 | 1, 53 | | 0.55 | |
|  | Condition | | 2.5 | 9, 486 | | | | **0.007*** | | | 1.2 | 9, 486 | | 0.30 | |
|  | Condition x Group | | 0.7 | 9, 486 | | | | 0.74 | | | 0.3 | 9, 486 | | 0.97 | |
| Superior frontal gyrus | Group | | 0.6 | 1, 54 | | | | 0.45 | | | 0.5 | 1, 54 | | 0.47 | |
|  | Condition | | 2.7 | 9, 486 | | | | **0.004*** | | | 3.4 | 9, 486 | | **< 0.001*** | |
|  | Condition x Group | | 2.0 | 9, 486 | | | | **0.036*** | | | 1.1 | 9, 486 | | 0.38 | |
| Middle temporal gyrus | Group | | 0.3 | | 1, 54 | | 0.59 | | | |  | | | | |
|  | Condition | | 3.5 | | 9, 486 | | **< 0.001*** | | | |  |  |  |  |  |
|  | Condition x Group | | 1.0 | | 9, 486 | | 0.43 | | | |  |  |  |  |  |

**Table S3. Fear generalization fMRI results (percent signal change) by group and morph**

| **Left Hemisphere** | | | | **Right Hemisphere** | | |
| --- | --- | --- | --- | --- | --- | --- |
|  | **CHR-** | **CHR+** |  | **CHR-** | **CHR+** |  |
|  | **Mean ± SE** | **Mean ± SE** | **p-value** | **Mean ± SE** | **Mean ± SE** | **p-value** |
| **Angular Gyrus** | | | | | | |
| CS+ | -0.15 ± 0.05 | -0.17 ± 0.09 | 0.86 | -0.10 ± 0.05 | -0.14 ± 0.08 | 0.71 |
| M1 | -0.27 ± 0.04 | -0.26 ± 0.05 | 0.85 | -0.21 ± 0.05 | -0.20 ± 0.05 | 0.91 |
| M2 | -0.19 ± 0.05 | -0.19 ± 0.05 | 0.99 | -0.17 ± 0.04 | -0.21 ± 0.05 | 0.64 |
| M3 | -0.19 ± 0.04 | -0.13 ± 0.05 | 0.43 | -0.18 ± 0.03 | -0.11 ± 0.05 | 0.27 |
| M4 | -0.20 ± 0.06 | -0.09 ± 0.05 | 0.33 | -0.19 ± 0.07 | -0.11 ± 0.05 | 0.50 |
| M5 | -0.18 ± 0.03 | -0.22 ± 0.06 | 0.49 | -0.19 ± 0.04 | -0.14 ± 0.05 | 0.44 |
| M6 | -0.13 ± 0.05 | -0.14 ± 0.04 | 0.94 | -0.19 ± 0.07 | -0.12 ± 0.05 | 0.59 |
| M7 | -0.11 ± 0.04 | -0.11 ± 0.06 | 0.99 | -0.11 ± 0.05 | -0.09 ± 0.06 | 0.82 |
| M8 | -0.17 ± 0.05 | -0.06 ± 0.05 | 0.16 | -0.15 ± 0.04 | -0.04 ± 0.05 | 0.13 |
| CS- | -0.14 ± 0.05 | -0.11 ± 0.07 | 0.69 | -0.16 ± 0.05 | -0.06 ± 0.04 | 0.29 |
| **Middle Temporal Gyrus** | | | | | | |
| CS+ | -0.13 ± 0.06 | -0.07 ± 0.11 | 0.63 |  |  |  |
| M1 | -0.32 ± 0.05 | -0.31 ± 0.11 | 0.92 |  |  |  |
| M2 | -0.27 ± 0.06 | -0.26 ± 0.07 | 0.92 |  |  |  |
| M3 | -0.28 ± 0.04 | -0.32 ± 0.10 | 0.68 |  |  |  |
| M4 | -0.30 ± 0.06 | -0.19 ± 0.06 | 0.30 |  |  |  |
| M5 | -0.19 ± 0.03 | -0.16 ± 0.07 | 0.62 |  |  |  |
| M6 | -0.16 ± 0.05 | -0.21 ± 0.08 | 0.58 |  |  |  |
| M7 | -0.10 ± 0.05 | -0.16 ± 0.08 | 0.54 |  |  |  |
| M8 | -0.13 ± 0.05 | -0.13 ± 0.09 | 1.00 |  |  |  |
| CS- | -0.22 ± 0.06 | 0.00 ± 0.08 | 0.05 |  |  |  |
| **Anterior Insula** | | | | | | |
| CS+ | 0.56 ± 0.06 | 0.16 ± 0.08 | **<0.01*** | 0.43 ± 0.06 | 0.41 ± 0.09 | 0.80 |
| M1 | 0.37 ± 0.06 | 0.26 ± 0.05 | 0.29 | 0.32 ± 0.04 | 0.46 ± 0.05 | 0.07 |
| M2 | 0.32 ± 0.06 | 0.28 ± 0.04 | 0.72 | 0.29 ± 0.06 | 0.50 ± 0.06 | 0.05 |
| M3 | 0.34 ± 0.07 | 0.16 ± 0.08 | 0.15 | 0.31 ± 0.05 | 0.34 ± 0.08 | 0.72 |
| M4 | 0.31 ± 0.04 | 0.14 ± 0.08 | 0.06 | 0.30 ± 0.05 | 0.27 ± 0.09 | 0.77 |
| M5 | 0.20 ± 0.05 | 0.13 ± 0.08 | 0.47 | 0.17 ± 0.05 | 0.31 ± 0.09 | 0.13 |
| M6 | 0.10 ± 0.05 | 0.20 ± 0.06 | 0.26 | 0.12 ± 0.05 | 0.25 ± 0.05 | 0.14 |
| M7 | 0.12 ± 0.05 | 0.02 ± 0.08 | 0.33 | 0.10 ± 0.06 | 0.15 ± 0.07 | 0.66 |
| M8 | 0.17 ± 0.04 | 0.19 ± 0.08 | 0.76 | 0.17 ± 0.05 | 0.27 ± 0.07 | 0.28 |
| CS- | 0.06 ± 0.05 | 0.07 ± 0.05 | 0.90 | -0.01 ± 0.07 | 0.20 ± 0.06 | 0.08 |
| **Posterior Cingulate Cortex** | | | | | | |
| CS+ | -0.14 ± 0.07 | -0.19 ± 0.07 | 0.67 | -0.04 ± 0.06 | -0.09 ± 0.07 | 0.63 |
| M1 | -0.30 ± 0.06 | -0.30 ± 0.07 | 0.99 | -0.16 ± 0.06 | -0.13 ± 0.05 | 0.75 |
| M2 | -0.22 ± 0.06 | -0.22 ± 0.07 | 0.98 | -0.12 ± 0.05 | -0.10 ± 0.05 | 0.80 |
| M3 | -0.27 ± 0.05 | -0.18 ± 0.06 | 0.30 | -0.16 ± 0.04 | -0.10 ± 0.06 | 0.39 |
| M4 | -0.20 ± 0.08 | -0.05 ± 0.06 | 0.28 | -0.09 ± 0.07 | -0.06 ± 0.05 | 0.76 |
| M5 | -0.14 ± 0.04 | -0.20 ± 0.05 | 0.38 | -0.08 ± 0.03 | -0.06 ± 0.03 | 0.78 |
| M6 | -0.18 ± 0.06 | -0.12 ± 0.07 | 0.63 | -0.09 ± 0.07 | -0.05 ± 0.08 | 0.72 |
| M7 | -0.10 ± 0.06 | -0.11 ± 0.07 | 0.93 | -0.06 ± 0.04 | 0.04 ± 0.06 | 0.23 |
| M8 | -0.09 ± 0.05 | -0.09 ± 0.05 | 0.99 | -0.07 ± 0.04 | -0.04 ± 0.04 | 0.65 |
| CS- | -0.17 ± 0.06 | -0.07 ± 0.05 | 0.30 | -0.10 ± 0.06 | -0.01 ± 0.05 | 0.39 |
| Superior Frontal Gyrus | | | | | | |
| CS+ | 0.27 ± 0.04 | 0.15 ± 0.07 | 0.12 | 0.36 ± 0.05 | 0.27 ± 0.08 | 0.34 |
| M1 | 0.09 ± 0.03 | 0.12 ± 0.05 | 0.67 | 0.22 ± 0.03 | 0.28 ± 0.06 | 0.40 |
| M2 | 0.07 ± 0.03 | 0.15 ± 0.05 | 0.17 | 0.21 ± 0.04 | 0.27 ± 0.06 | 0.41 |
| M3 | 0.10 ± 0.03 | 0.05 ± 0.04 | 0.36 | 0.21 ± 0.04 | 0.20 ± 0.07 | 0.99 |
| M4 | 0.05 ± 0.05 | 0.09 ± 0.04 | 0.60 | 0.17 ± 0.06 | 0.26 ± 0.06 | 0.42 |
| M5 | 0.07 ± 0.03 | 0.12 ± 0.04 | 0.34 | 0.17 ± 0.03 | 0.21 ± 0.05 | 0.51 |
| M6 | 0.05 ± 0.03 | 0.12 ± 0.04 | 0.23 | 0.19 ± 0.04 | 0.19 ± 0.05 | 0.94 |
| M7 | 0.06 ± 0.02 | 0.08 ± 0.02 | 0.66 | 0.11 ± 0.03 | 0.14 ± 0.05 | 0.55 |
| M8 | 0.10 ± 0.04 | 0.13 ± 0.07 | 0.64 | 0.15 ± 0.05 | 0.21 ± 0.08 | 0.52 |
| CS- | 0.03 ± 0.03 | 0.17 ± 0.05 | **0.03*** | 0.08 ± 0.04 | 0.22 ± 0.06 | 0.07 |

* p < 0.05

**Table S4. Correlations across the CHR+ Sample**

|  | Negative symptoms | Anxiety | Ratings function threshold | Ratings function max | Left insula function threshold | Left insula function max |
| --- | --- | --- | --- | --- | --- | --- |
| Positive symptoms | **0.48** | 0.15 | 0.09 | -0.06 | -0.16 | 0.22 |
|  | **0.03*** | 0.5 | 0.69 | 0.79 | 0.56 | 0.41 |
| Negative symptoms |  | 0.2 | 0.14 | **-0.44** | 0.18 | 0.05 |
|  |  | 0.36 | 0.54 | **0.04*** | 0.51 | 0.86 |
| Anxiety |  |  | 0.3 | 0 | -0.03 | 0 |
|  |  |  | 0.17 | 1 | 0.91 | 0.99 |
| Ratings function  threshold |  |  |  | -0.3 | 0.51 | -0.15 |
|  |  |  |  | 0.17 | 0.04* | 0.58 |
| Ratings function  max |  |  |  |  | -0.22 | -0.09 |
|  |  |  |  |  | 0.41 | 0.73 |
| Left Insula function threshold |  |  |  |  |  | **-0.71** |
|  |  |  |  |  |  | **0.00*** |

* p < 0.05

**Table S5. Correlations across the full sample.**

|  | Negative Symptoms | Anxiety | Ratings function threshold | Ratings function max | Left insula function threshold | Left insula function max |
| --- | --- | --- | --- | --- | --- | --- |
| Positive Symptoms | **0.56** | 0.49 | 0.1 | -0.06 | 0.23 | **-0.36** |
|  | **0.00*** | 0.00* | 0.42 | 0.64 | 0.08 | **0.01*** |
| Negative Symptoms |  | 0.37 | 0.13 | **-0.31** | 0.32 | **-0.28** |
|  |  | 0.00* | 0.29 | **0.01*** | 0.02* | **0.04*** |
| Anxiety |  |  | 0.28 | -0.03 | **0.28** | **-0.35** |
|  |  |  | 0.02* | 0.83 | **0.03*** | **0.01*** |
| Ratings function threshold |  |  |  | -0.28 | 0 | 0.02 |
|  |  |  |  | 0.02* | 0.98 | 0.87 |
| Ratings function max |  |  |  |  | -0.01 | -0.05 |
|  |  |  |  |  | 0.93 | 0.73 |
| Left insula function threshold |  |  |  |  |  | **-0.83** |
|  |  |  |  |  |  | **0.00*** |

* p < 0.05
